# Risk Factors for Early PICC Removal: A Retrospective Study of Adult Inpatients at an Academic Medical Center

**DOI:** 10.1101/2022.02.07.22270642

**Authors:** Burton H. Shen, Lindsey Mahoney, Janine Molino, Leonard Mermel

## Abstract

**Background:** Use of PICCs has been rising since 2001. They are used when long-term intravenous access is needed and for blood draws in patients with difficult venous access.

**Objective:** To determine which risk factors contribute to inappropriate PICC line insertion defined as removal of a PICC within five days of insertion for reasons other than a PICC complication.

**Design:** Retrospective, observational study.

**Setting:** Tertiary-care, Level 1 trauma center.

**Patients:** Adult patients with a PICC removed 1/1/2017 to 5/4/2020.

**Measurements:** Frequency of PICC removal within five days of insertion and associated risk factors for early removal.

**Results:** Between 1/1/17 and 5/4/2020, 995 of 5348 PICCs inserted by the IV nursing team were removed within five days (19%). In 2017, 5 of 429 PICCs developed a central line-associated infection (1.2%) and 29 of 429 PICCs developed symptomatic venous thromboembolism (6.7%). Patients with PICCs whose primary service was a medical subspecialty were independently at higher risk of early removal (OR 1.44, 95% CI 1.14, 1.83); weekday insertion was independently associated with a lower likelihood of early removal compared to weekend insertion (OR 0.60; 95% CI 0.49, 0.75).

**Limitation:** PICC removal after discharge was not assessed and paper records were likely incomplete and biased.

**Conclusion:** Nearly one in five PICCs were removed within five days. Patients whose primary team was a medical subspecialty were at independently higher risk of early removal.

## Introduction

Peripherally-inserted central catheters (PICCs) are an important component of medical care.(1, 2) They are frequently utilized when long-term intravenous (IV) access is required for antibiotic administration, medications requiring central venous access, parenteral nutrition, chemotherapy, or frequent blood draws in patients with difficult venous access.(3) Use of PICCs has been rising since 2001 due to their increased availability, ease of insertion and safety compared with central venous catheters (CVCs) inserted at other sites, and durability over extended periods of time;(1, 4, 5) however, the average dwell time for PICCs has been decreasing.(5)

In hospitalized patients, insertion of a PICC for infusion of peripherally-compatible fluids, frequent phlebotomy, or difficult venous access is considered inappropriate if the expected duration of catheterization is five days or less.(2) However, infusion of irritants or vesicants such as parenteral nutrition or chemotherapy is appropriate for any proposed duration.(2) Two prospective, multi-center studies found that approximately 25% of PICCs in hospitalized patients had dwell times of five days or less.(3, 6) Known complications from PICCs include venous thromboembolism (VTE), central line-associated bloodstream infection (CLABSI), exit site infection, catheter lumen occlusion, and catheter tip migration, all of which are a cause of significant morbidity, potential mortality, and increased healthcare cost.(7-11)

Mitigating inappropriate, short-term use of PICCs has the potential for harm reduction and cost-savings. The goals of this study were to assess: frequency and risk factors associated with catheter dwell time of five or fewer days; complications resulting from PICC use and determine if these complications were associated with the number of catheter lumens; and to assess changes in PICC dwell time during the study period.

## Methods

### Study Design

We performed a retrospective, observational study at Rhode Island Hospital, a tertiary-care, Level 1 trauma center licensed for 719 beds. Data were collected from two sources: paper records collected by our IV nursing team and the electronic health record (EHR). The IV nursing team kept paper records of PICC insertions and removals through 12/31/2017. On 1/1/2018, the IV nursing team transitioned to full utilization of the EHR. Two reviewers (BHS, LM) used the paper records from 1/1/17 to 12/31/17 to identify any patient who had a PICC removed during their hospital stay. The correct patient and hospital encounter was then identified in the EHR. We identified patient demographics and characteristics including age, gender, race, and body mass index (BMI) at the time of PICC insertion. We identified the service team overseeing the patient’s care at the time the PICC was ordered. Possible care teams included internal medicine (teaching service), hospital medicine (non-teaching service), general surgery, orthopedic surgery, neurosurgery, other medicine subspecialty, or other surgical subspecialty service. Podiatry, otolaryngology, plastic surgery, orthopedic surgery, neurosurgery, and dental surgery are all separate training programs from other surgical subspecialities. Among these, orthopedics and neurosurgery are the only ones that regularly admit patients to their service at our hospital; the other services admit to the medicine service for co-management.

We also identified characteristics of each PICC including number of lumens, indications for insertion and removal, order date, and time of insertion and removal. Lastly, we identified any PICC-associated complications. Symptomatic venous thromboembolism (VTE) of the extremity used for PICC placement and CLABSI were considered major complications. Symptomatic VTE was defined as any patient with a PICC who developed swelling, redness or pain that prompted imaging and which confirmed a thrombus. For CLABSI, we used the CDC National Healthcare Safety Network (CDC/NHSN) definition.(12) Minor complications included catheter occlusion, superficial thrombosis, mechanical complications such as kinking or coiling of the catheter, exit site infection, or catheter tip migration. All the information collected was recorded and stored in a REDCap database. Data from 1/1/2018 to 5/4/2020 was obtained exclusively from the EHR. Data collected during this time included patient demographic information and objective PICC data including number of lumens, order date and time, insertion date and time, and removal date and time. We were unable to obtain the service team at the time of insertion, indications for PICC insertion, or complications related to the PICC since this free text data is difficult to obtain from an automated data pull. An EHR automated data pull was also obtained for the January 2017-December 2017 time period to compare with and validate the manually collected data. The Lifespan Institutional Review Board approved this project.

### Inclusion and Exclusion Criteria

Inpatients at least 18 years of age who had a PICC inserted by our IV nursing team on or after 1/1/2017 and removed before 5/4/2020 were included in this study. Patients were excluded if their PICC was placed at an outside hospital or by other services (e.g., interventional radiology), as these PICCs were not included in the paper records kept by the IV nursing team. PICCs removed due to complications were not excluded.

### Statistical Analysis

Data were imported into SAS version 9.4 (SAS Institute Inc., Cary, NC) for data management and hypothesis testing. The assessment of complications related to PICC use was based on PICCs inserted on or after 1/1/17 and removed before 12/31/17 collected from paper records. The assessment of risk factors for dwell time of five days or fewer was based on EHR data of PICCs inserted on or after 1/1/17 and removed before 5/4/20. Descriptive statistics were obtained for the study sample characteristics. Mean and standard deviation were reported for the continuous variables while frequency and percentage were reported for the binary and categorical variables. The prevalence of PICCs placed that were removed within 5 days, as well as the prevalence of major and minor PICC complications, were reported. Generalized estimating equations (GEE) were used (1) to examine the factors associated with PICCs removed within five days of insertion (GEE with a binomial distribution and logit link); (2) to examine which PICC complications were associated with the number of lumens (GEE with a negative binomial distribution and logit link); and (3) to examine whether the rate of early removal changed over time (GEE with a binomial distribution and logit link). Classical sandwich estimators were used to protect against possible model misspecification. A p-value < 0.05 was used to determine statistical significance.

## Results

Approximately one-third of PICCs were placed in patients on a medical subspecialty service and over half of the catheters were double lumen PICCs (Table 1).

**Table 1:**
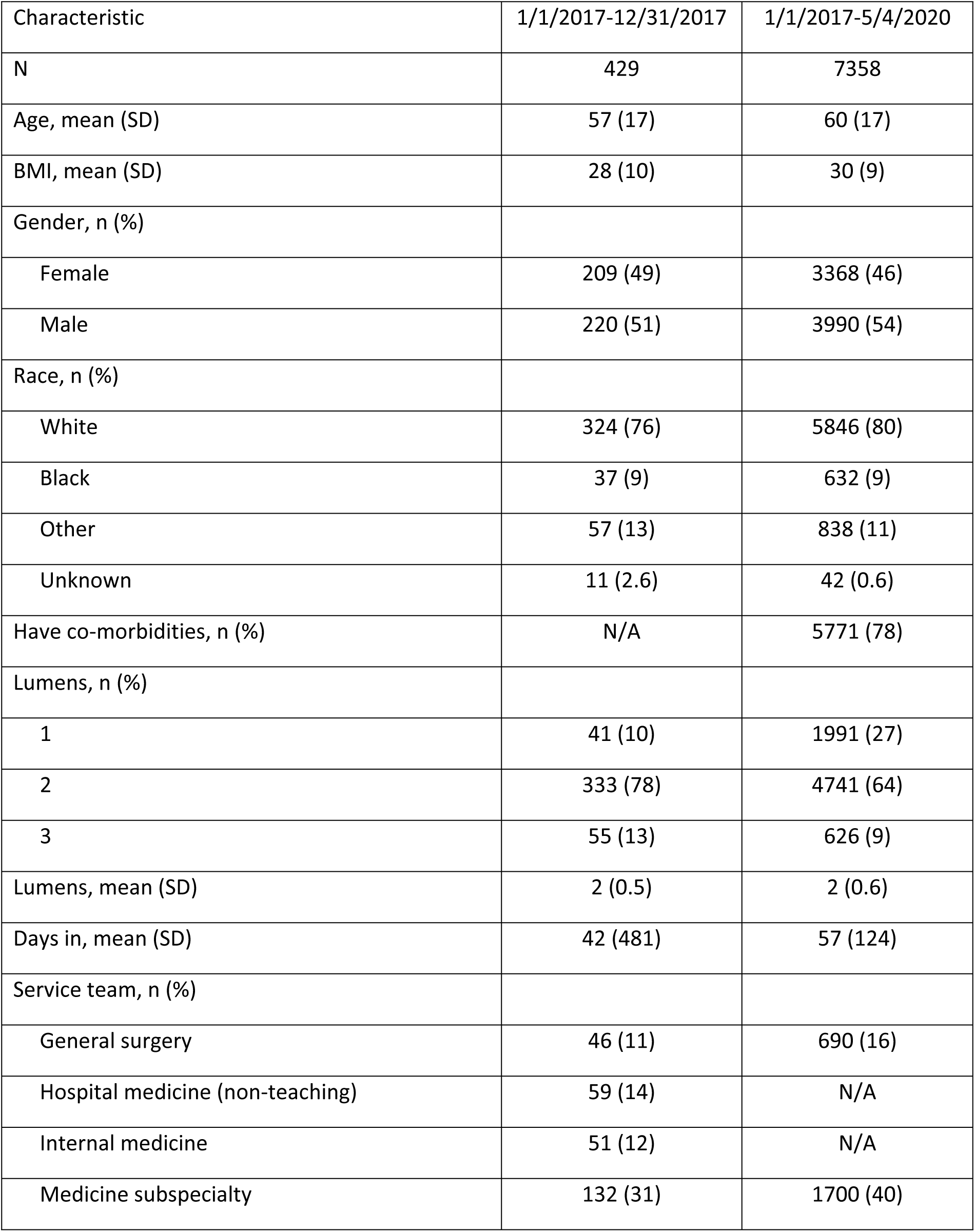
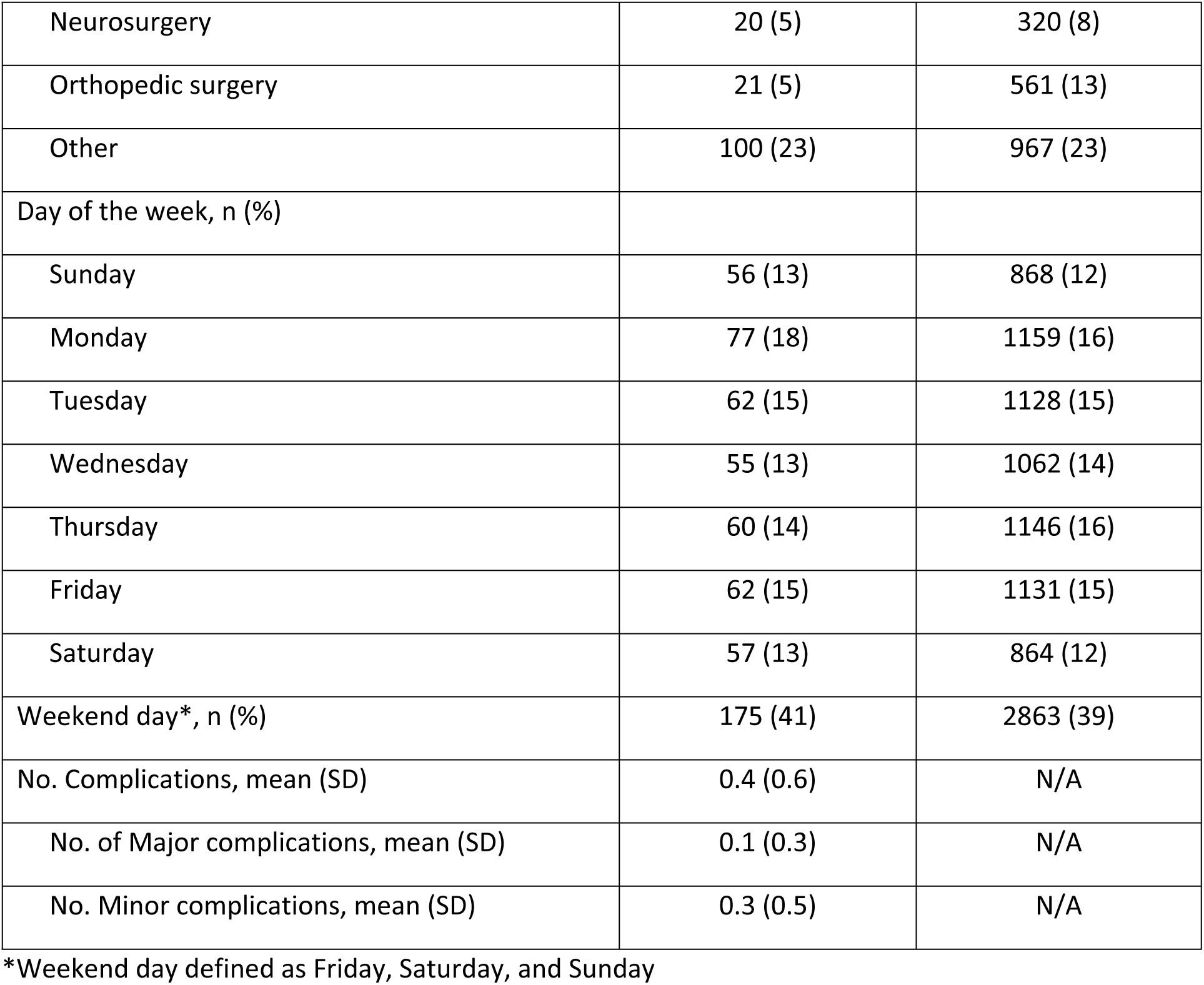
Characteristics of patients and PICCs

Female gender was associated with early removal in three of the four models. From 1/1/17 through 5/4/20, 995 of 5348 PICCs were removed within five days (19%, Table 2).

**Table 2:** Outcomes of patients with PICCs

| Outcome | 1/1/2017-12/31/2017 | 1/1/2017-5/4/2020 |
| --- | --- | --- |
| PICC removal within 5 days of insertion | 141/ 429 (33%) | 995/5348 (19%)* |
| Symptomatic venous thromboembolism | 29/ 429 (7%) | N/A |
| Central line-associated bloodstream infection | 5/ 429 (1%) | N/A |
| Catheter occlusion | 77/ 429 (18%) | N/A |
| Superficial venous thrombosis | 7/ 429 (2%) | N/A |
| Mechanical complication | 13/ 429 (3%) | N/A |
| Exit site infection | 0/ 429 (0%) | N/A |
| Catheter tip migration | 44/ 429 (10%) | N/A |
| Number without major or minor PICC complications | 303/ 429 (71%) | N/A |

Note: \*Missing data for 2010 patients due to no PICC removal date in the EHR

Medical subspecialties were independently associated with a higher likelihood of early PICC removal (OR 1.44, 95% CI 1.14, 1.83), while weekday insertion (Monday through Thursday) was independently associated with a lower likelihood of early removal (OR 0.60; 95% CI 0.49, 0.75, Table 3). Complications for all PICCs inserted were not associated with the number of PICC lumens (Table 4).

**Table 3:** Potential risk factors associated with early PICC removal by multivariable analysis

| Characteristic | OR (95% CI) | P |
| --- | --- | --- |
| Female | 1.16 (0.96, 1.40) | 0.13 |
| Age |  |  |
| BMI | 1.01 (0.99, 1.02) | 0.39 |
| White |  |  |
| Co-morbidity |  |  |
| Team |  | <.001 |
| General surgery | 0.95 (0.71, 1.28) |  |
| Hospital medicine | N/A |  |
| Internal medicine | N/A |  |
| Med subspecialty | 1.44 (1.14, 1.83) |  |
| Neurosurgery | 0.96 (0.63, 1.45) |  |
| Orthopedic surgery | 0.66 (0.43, 1.01) |  |
| Other | *Reference* |  |
| PICC lumens | 1.00 (0.84, 1.20) | 0.97 |
| Indication |  |  |
| Antibiotics | N/A |  |
| Chemotherapy | N/A |  |
| Difficult venous access | N/A |  |
| Long term access | N/A |  |
| Meds requiring central access | N/A |  |
| Incompatible fluids | N/A |  |
| Parenteral nutrition | N/A |  |
| Unknown | N/A |  |
| Multiple | N/A |  |
| No. complications | N/A |  |
| Day of week |  | <0.001 |
| Sunday | *Reference* |  |
| Monday | 0.65 (0.46, 0.91) |  |
| Tuesday | 0.60 (0.42, 0.85) |  |
| Wednesday | 0.52 (0.36, 0.75) |  |
| Thursday | 0.62 (0.44, 0.88) |  |
| Friday | 0.56 (0.39, 0.79) |  |
| Saturday | 0.96 (0.67, 1.36) |  |

**Table 4:** Complications associated with number of PICC lumens in the 2017 Manual Chart Review Data*

| Complication | IRR | 95% CI | P |
| --- | --- | --- | --- |
| Symptomatic VTE | 1.02 | (0.94, 1.11) | 0.66 |
| CLABSI | 0.88 | (0.73, 1.08) | 0.24 |
| Catheter occlusion | 1.04 | (0.99, 1.08) | 0.14 |
| Catheter tip migration | 0.99 | (0.92, 1.08) | 0.90 |
| No. complications | 1.02 | (0.98,1.05) | 0.35 |

\*Based on univariable GEEs and lumens treated as a count (i.e., 1, 2, or 3 PICC lumens)

## Discussion

Nearly one in five PICCs were removed within five days, similar to prior studies. (3) Patients whose primary team was a medical subspecialty such as hematology-oncology or critical care, were at greater risk of having early PICC removal. This is not unexpected, as a PICC may be used for chemotherapy for fewer than five days, or for critical care of a patient whose condition improves within five days of insertion. Approximately one in three PICC removals were on medical subspecialty services, though we did not separate which were on critical care or hematology-oncology services. We found that dwell times may differ based on day of the week, or whether it is a weekday or weekend. When the order for PICC insertion occurred on a weekday, there was a significantly lower likelihood of early PICC removal. This may reflect PICC insertion during the weekend when there was limited phlebotomy services and nursing assistance and PICC removal the following weekdays when services for peripheral IV placement, phlebotomy are more available.

We could not confirm that the number of PICC lumens is associated with increased risk of complications such as CLABSI; however, our study may have been underpowered to assess for these outcome measures.(13)

Data for this study was shared with the hospital administration leading to a change in the name of the IV team to the vascular access team and a change in hospital policy such that practitioners were no longer able to order PICC for insertion by the team. Instead, providers were able to order a vascular access consult so the team could assess the patient based on information provided and medical record review to make the best decision regarding the most appropriate vascular access for the patient. We are tracking vascular access to assess the impact of these changes in hopes of reducing early PICC removal and improving patient outcomes.

Our study has a number of limitations. The paper records in 2017 were incomplete due to user omission, absent medical record numbers, and difficulty with handwriting. Additionally, there is likely selection bias regarding which PICC removals were documented. It is unclear what direction the selection bias may lean towards, as there are myriad reasons why documenting could have been variable on any given day. Since the paper records did not include all patients whose PICC was removed, complications may be over or under represented. We only assessed PICCs that were removed during hospitalization. Thus, we did not asses the many PICCs removed from patients after hospital discharge.

## Data Availability

All relevant data are within the manuscript and its Supporting Information files.

